# Screening COVID-19 by Swaasa AI Platform using cough sounds: A cross-sectional study

**DOI:** 10.1101/2022.11.02.22281821

**Authors:** P Padmalatha, Gowrisree Rudraraju, Narayana Rao Sripada, Baswaraj Mamidgi, Charishma Gottipulla, Charan Jalukuru, ShubhaDeepti Palreddy, Nikhil kumar Reddy Bhoge, Priyanka Firmal, Venkat Yechuri, PV Sudhakar, B Devimadhavi, S Srinivas, K K L Prasad, Niranjan Joshi

**Affiliations:** Andhra Medical College, Visakhapatnam, India; Salcit Technologies, Jayabheri Silicon Towers, Hyderabad India; C-CAMP; Guntur Medical College, India

**Keywords:** COVID-19, Cough signature, Convolutional Neural Network (CNN), Tabular model, Machine learning

## Abstract

The Advent of Artificial Intelligence (AI) has led to the use of auditory data for detecting various diseases, including COVID-19. SARS-CoV-2 infection has claimed more than 6 million lives till date and hence, needs a robust screening technique to control the disease spread. In the present study we developed and validated the Swaasa AI platform for screening and prioritizing COVID-19 patients based on the signature cough sound and the symptoms presented by the subjects. The cough data records collected from 234 COVID-19 suspects were subjected to validate the convolutional neural network (CNN) architecture and tabular features-based algorithm. The likelihood of the disease was predicted by combining the final output obtained from both the models. In the clinical validation phase, Swaasa was found to be 75.54% accurate in detecting the likely presence of COVID-19 with 95.45% sensitivity and 73.46% specificity. The pilot testing of Swaasa was carried out on 183 presumptive COVID subjects, out of which 82 subjects were found to be positive for the disease by Swaasa. Among them, 58 subjects were truly COVID-19 positive, which corresponds to a Positive Predictive Value of 70.73%. The currently available rapid screening methods are very costly and require technical expertise, therefore a cost effective, remote monitoring tool would be very beneficial for preliminary screening of the potential COVID-19 subject.

## Introduction

The SARS-CoV-2 infection first surfaced at the end of December 2019, affecting nearly 600 million people till now across the globe [1]. The virus transmission begins once a healthy individual is exposed to the respiratory droplets originated from an infected person. The average incubation period for the disease symptoms to manifest varies from 2-14 days [2]. The early symptoms comprise dry cough, fever, fatigue, loss of smell and taste. In a few cases the patient may experience shortness of breath, cardiac issues, and pneumonia like symptoms, which can ultimately result in death. Many people are also experiencing post-covid acute symptoms which affects their overall health status [3, 4]. The containment of COVID-19 outbreaks became very difficult because of the unavailability of quick and effective pre-screening techniques. Most of the viral and serological testing methods available are very expensive, time consuming, require technical expertise and are not always reliable, especially in detecting the new SARC-CoV-2 variants [5, 6].

Cough has been presented as a common symptom of COVID-19 [7], which is mainly responsible for removing any obstruction in the airways via explosive expulsion of the air [8]. Cough is a common decipher of various respiratory diseases, including tuberculosis and now SARC-CoV-2 infection, which results in the dissemination of airborne infectious aerosols into the environment [9, 10]. It has already been reported that a characteristic glottis movement is observed under specific diseased conditions, which can be utilized to differentiate the origin of coughs in different circumstances such as pertussis, bronchitis, and asthma [11, 12]. As cough is the main classifier of the presence of COVID-19, there are various reports which suggest that it could be used for mass screening of the disease [13, 14]. Still a thorough investigation is needed to make use of cough sound analysis as a determinant of the presence or absence of SARC-CoV-2 infection.

In the past few years, a substantial increase in the studies has been observed in terms of exploring Artificial Intelligence (AI)-based algorithms in the field of medicine, including the analysis of cough sound data for determining various respiratory diseases [15, 16]. Various groups have highlighted the importance of machine-learning techniques in detecting COVID-19 using cough as opposed to pre-screening methods such as, RT-PCR [17–21]. Despite so much of investigation for developing a desired AI-based tool, no such device is available in the market yet. The main factors being the acquisition of cough sound data from crowdsource open access datasets and lack of proper technical/clinical validations to scale up these tools for mass screening of COVID subjects [22, 23].

In the current study we have adopted a different approach. Our data comprises a good amount of COVID-19 positive coughs, coughs from healthy subjects and coughs from patients suffering with various respiratory conditions. We did carry feature analysis of COVID-19 and non-COVID-19 coughs. Our analysis shows that COVID-19 related cough has a unique signature, which can be identified by a machine learning model. Further we used a unique multimodal architecture based on CNN and tabular features. The coughs are converted to Mel-frequency cepstral coefficient spectrograms, the spectrogram images are fed as inputs to CNN classifier for classification. Simultaneously, a tabular model is trained using features extracted from frequency domain and time domain. The output from these two models is combined to detect the likely presence of COVID-19 (yes/no/inconclusive). We obtained a 96% accuracy on the test dataset in the derivation and 76% in the validation phase. Additionally, the tool has been thoroughly validated in the clinical settings and has proved to have a positive prediction value of 70.73% in the real time scenario. This remote tool is highly desired for rapid and cost-effective non-invasive screening of COVID-19 cases. However, a large-scale validation study is needed for improvising the accuracy of the tool for making it more accessible for diverse ethnicities located worldwide.

## Methods and Materials

### Sample size estimation and Data Collection

According to the sample size calculation, a total of 1152 comprising 40% COVID-19 positive cases and 60% control subjects was appropriate for validating if the device could detect COVID-19 with a 90% sensitivity on considering a 2.5% error for a 95% confidence interval (CI) and a prevalence of 0.75%. Considering all the conditions, we pooled the data collected from four individual clinical trial studies for developing and evaluating our model. We considered a total of 1052 participants in the present study, out of which 62% were controls. Control subjects comprise healthy individuals as well as subjects who were displaying various respiratory disease symptoms and came out positive for conditions such as asthma, Chronic Obstructive Pulmonary Disease (COPD), Interstitial lung disease (ILD), pneumonia but were negative for COVID-19 via RT-PCR.

The cough data was collected at Andhra Medical College, Visakhapatnam, India as a part of individual studies entitled “Development, Validation, Pilot Deployment of an ultra-scalable technology - Swaasa AI, as an auxiliary to COVID-19 Rapid test”, “COVID-19 Cough Sound Analysis Using Swaasa Artificial Intelligence Platform”. The studies were registered under Clinical Trials Registry-India CTRI/2021/09/036489, CTRI/2021/07/035096, and were begun after getting the approval from the AMC-Institutional Ethics Committee (IEC). The methodologies performed throughout the study were in accordance with the set guidelines. A duly signed written informed consent was also collected from all the enrolled subjects before starting the trial, which was followed by collection of the demographic details and the vital signs. Patients were then interviewed for the Part I of the St. George’s Respiratory Questionnaire (SGRQ) and COVID-19 symptoms in order to gather their symptoms [24]. Next, the cough sound was collected by a trained health care personnel. Each subject recorded multiple coughs (3-4 times), taking a breath in between every 15 seconds record interval. Following the cough sample collection, the patients were subjected to a reference standard test (RT-PCR).

The inclusion criteria for the enrolment are that the Patients must be (a) male and female patients age ≥ 18 years, who were (b) recently diagnosed with COVID 19 (for validation phase only) and were (c) able to read, understand and sign the informed consent form. Whereas male and female patients age < 18 years, who were (b) on ventilators support, (c) asymptomatic patients attending isolation ward for COVID testing and (d) Pregnant females were completely excluded from the current study. COVID precautionary and infection control measures were followed strictly.

### Study design and Patient recruitment

The cross-sectional studies were undertaken in three phases. In the derivation phase (Phase 1) multiple data points (cough recordings) were collected from every subject to develop a robust model. Whereas, in the Phase 2 (validation) and Phase 3 (pilot test) only one data point (one cough recording) was collected.

**Phase 1**: Derivation with an objective to quantify the technical / analytical performance of the device by establishing a unique cough signature for COVID-19.

Sample size: For derivation phase, a total of 642 subjects were considered, which were part of two separate derivation studies. Out of which 252 subjects were tested positive for COVID-19 by RT-PCR. Figure 1 shows the age and gender distribution of the recruited subjects.

**Figure 1:**
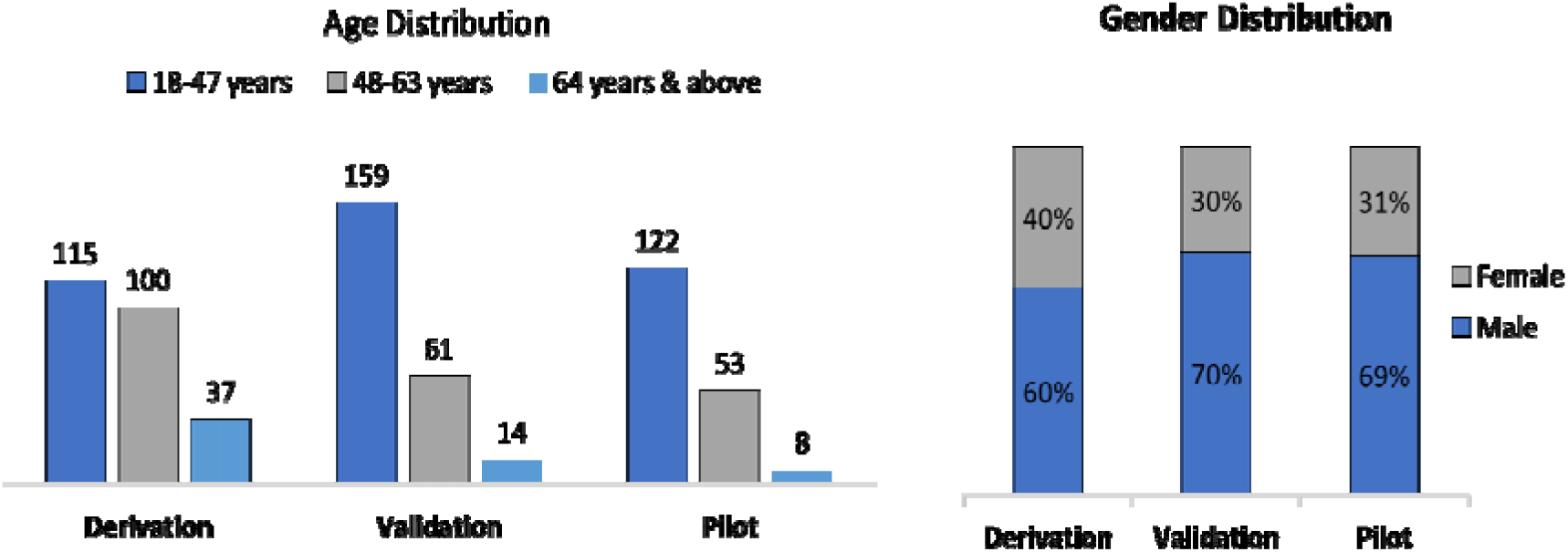
Data distribution in the derivation phase, validation phase and pilot testing.

**Phase 2**: Clinical validation to quantify the performance of the device against clinical diagnosis based on reference standard test.

Sample size: For validation, 234 subjects were recruited from various locations including, isolation ward, COVID testing centre-King George Hospital, Visakhapatnam, COVID testing centre-GHCCD, Visakhapatnam and were subjected to the screening test using Swaasa AI Platform, as well as to the standard reference testing for diagnosis of COVID-19 i.e., RT-PCR. We compared the results obtained from the Swaasa AI Platform to the standard diagnostic testing.

**Phase 3**: Pilot test to quantify the effectiveness of the device when deployed as a screening tool prior to diagnosis.

Sample size: In pilot deployment, we enrolled 183 presumptive COVID-19 cases from a peripheral health care centre, RHC Simhachalam.

### Data analysis

For phase 1 and phase 2, Swaasa’s performance was compared with diagnosis based on RT-PCR test. A consolidated test summary sheet was generated, which contained the results obtained from the classical gold standard diagnosis methods along with the Swaasa output. A statistician then compared both the results. The results obtained from the Swaasa AI platform were not accessible to the Physicians at any stage. For phase 3 the effectiveness of Swaasa was measured using the ratio of patients truly diagnosed as positive to all those who had positive test results based on Swaasa.

### Event Extraction

Moving windowed signal standard deviation technique was used for extracting the events from the collected cough records [25]. A cough/non-cough classifier was used to separate the cough recordings collected from 252 COVID-19 positive subjects during the derivation phase into true cough and non-cough events like speech, silence, fan sounds, vehicle horns, and noise. A total of 1946 cough events were extracted at this step from both COVID positive and negative subjects.

### Feature Extraction

Both the temporal and frequency domains of each cough event were used to extract the features. Zero crossing rate (ZCR) and energy are the two critical time domain features that were considered. Mel-frequency cepstrum (MFC), Spectral centroid, Spectral bandwidth, and Spectral roll off were the frequency domain features that were used for the data analysis [22].

The features were extracted from each frame of the cough signal. The average frame time was typically 20 milliseconds long. The coughing period can last anywhere from 200 to 700 milliseconds.

### Prediction Model

Coughs were converted to Mel-frequency cepstral coefficient (MFCC) spectrograms to construct a multimodal architecture based on Convolutional Neural Network (CNN) and tabular features. Spectrogram images were fed as input to a CNN classifier for classification. At the same time, a tabular model was trained using features extracted from the frequency and time domains. Feature selection techniques have been used to remove redundant features and to identify the key features.

The total features extracted were 209, that includes age, gender, 120 MFCC (40 MFCC, 40 first derivatives of MFCC, 40 second order derivatives of MFCC), 9 spectral features (spectral centroid, spectral roll-off, spectral bandwidth, dominant frequency, spectral skewness, spectral kurtosis, spectral crest, spectral spread and spectral entropy), 33 chroma features (11 chroma, 11 first derivatives of chroma, 11 second derivatives of chroma), 18 contrast features (6 contrast, 6 first derivatives of contrast, 6 second derivatives of contrast), 15 tonnentz features (5 tonnentz, 5 first derivatives of tonnentz, 5 second derivatives of tonnentz), 3 Zero-crossing rate (ZCR, first derivatives of ZCR, second derivatives of ZCR), 3 Energy (Energy, first derivatives of energy, second derivatives of energy), 3 skewness (skewness, first derivatives of skewness, second derivatives of skewness), 3 kurtosis (kurtosis, first derivatives of kurtosis, second derivatives of kurtosis). We performed correlation analysis and recursive feature elimination (RFE) on these traits to rank them according to importance. As part of feature selection, we removed irrelevant features and reduced the number of features in the tabular model to 170.

The output of CNN and tabular models was merged to detect the presence of COVID-19 (yes/no/inconclusive). When the model is unsure if COVID-19 will be detected as yes/no, it provides an inconclusive output as shown in the block diagram Figure 2.

**Figure 2:**
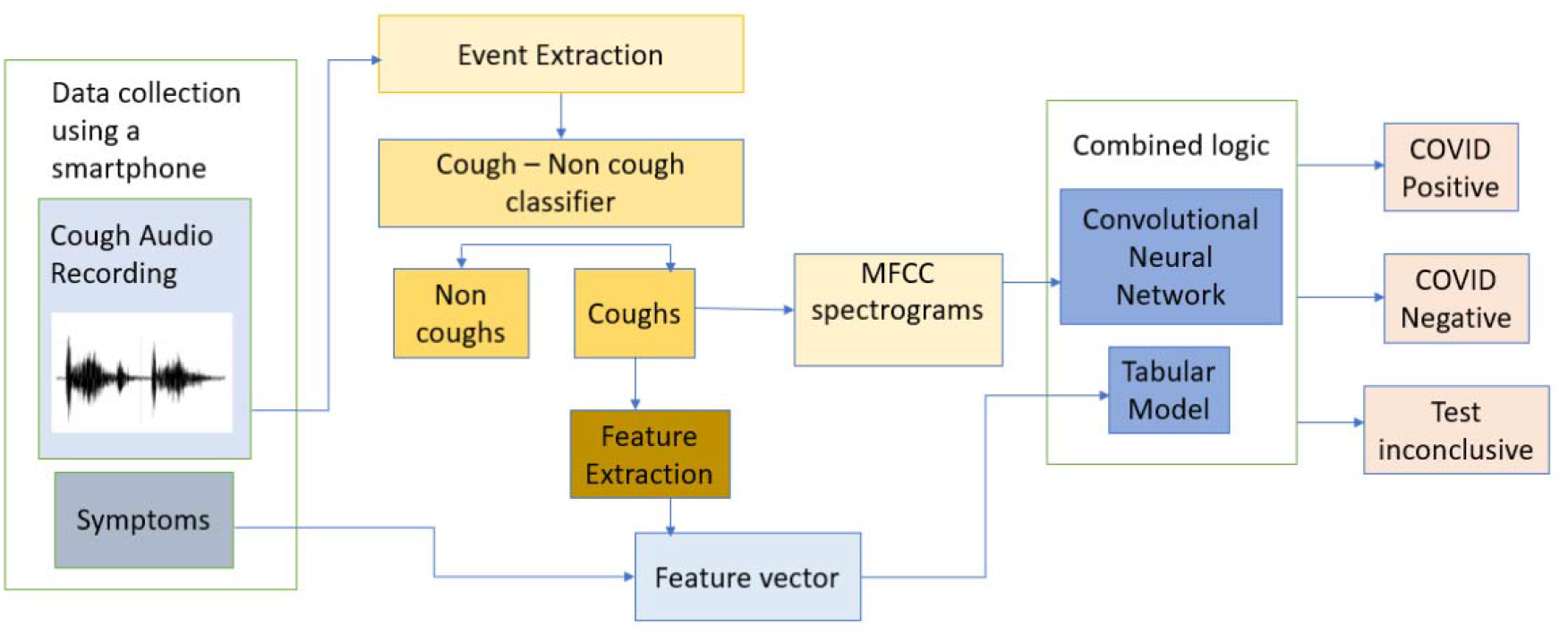
Block Diagram illustrating the flow of the COVID-19 prediction model.

Spectrograms were given as input to a CNN (convolutional neural network) that was pretrained (using transfer learning (Resnet34) with Imagenet for training. In parallel, a tabular model with primary and secondary characteristics was trained. From each of these two models, the last fully connected layer was removed, combined with a new fully connected layer (connected layer), again passed through the linear layer, the activation layer of the final output layer. We call this approach of combining the last layers of two models as combined logic.

### LIME Representation

The green portion of the Local interpretable model-agnostic explanations (LIME) representation [27] illustrates instances in which the model responded positively to a given class, whereas the red portion highlights instances in which it responded negatively. To “explain” a prediction, we refer to the display of textual or visual artefacts that give qualitative understanding of the link between the instance’s components (such as words in text, patches in a picture, etc.) and the prediction made by the model.

### Statistical significance

A comprehensive assessment of model performance on the test set includes accuracy, sensitivity, specificity, positive predictive value (PPV), negative predictive value (NPV), and ROC. To measure the variability of these parameters, we used the Clopper-Pearson method [28] with 95% confidence intervals. To better understand the model’s performance in screening COVID-19 subjects, we also calculated the confusion matrix across the test set.

## Results

### Performance parameters in Model derivation phase

Cough sound data was collected from 252 COVID-19 positive subjects in the derivation phase. Data collected from 390 COVID-19 negative subjects in one of our earlier studies was also considered in this phase. Among 252 subjects, 60% were male and 40% were female, with age ranging from 18 years to 64 & above. Subjects were confirmed with COVID-19 by standard diagnosis methods. In this phase multiple data points were collected from the subjects. Each data point was called a record. A total of 803 cough records were collected from 252 patients.

The 252 subject data was divided into training (173) and test (79). The training data was internally divided into training and validation as required to build as well as optimize the model performance based on K-fold cross validation technique. All the 252 subject data was annotated with disease condition as COVID-19 i.e., COVID-19 likely as “yes”. For COVID unlikely, data representing other disease conditions was added from pre-existing datasets [29] (collected part of earlier studies) in various propositions. A total of 1213 records data was added to various classifiers. The final confusion matrix for derivation phase is represented in Table 1. The performance parameters such as accuracy, sensitivity, specificity, and AUC (area under the curve) of the model in the derivation phase are enlisted in Table 2.

**Table 1:** Final Confusion matrix for the derivation phase

|  | COVID-19 Likely - Yes | COVID-19 Likely - No |
| --- | --- | --- |
| COVID-19 Likely - Yes | 256 (TP) | 11 (FN) |
| COVID-19 Likely - No | 12 (FP) | 278 (TN) |

**Table 2:** Performance metrics of the derivation phase

|  |  |
| --- | --- |
| Accuracy | 96% |
| Sensitivity | 95.8% |
| Specificity | 95.6% |
| AUC | 0.95 |

The model was also evaluated on crowdsourced data, which includes COVID-19 Likely “yes” data from EPFL and Cambridge datasets, whereas COVID-19 Likely “no” is the in-house data. The final confusion matrix for this dataset is presented in Table 3. The performance parameters for the same are listed in Table 4. In both the cases an AUC value of > 0.85 AUC has been achieved.

**Table 3:** Final Confusion matrix for the Crowdsource (test) data during derivation phase

|  | COVID-19 Likely - Yes | COVID-19 Likely - No |
| --- | --- | --- |
| COVID-19 Likely - Yes | 527 (TP) | 91 (FN) |
| COVID-19 Likely - No | 57 (FP) | 443 (TN) |

**Table 4:** Performance metrics of Crowdsource (test) data during derivation phase

|  |  |
| --- | --- |
| **Accuracy** | 86% |
| **Sensitivity** | 85% |
| **Specificity** | 88% |
| **AUC** | 0.855 |

The features listed in Table 5 depicts the mean value of the features extracted from individual frames, where we have considered normal as well as respiratory diseases data other than COVID from our previous validation study conducted at Apollo Hospitals, Hyderabad

**Table 5:** Table showing mean values of the Zero crossing rate (ZCR), spectral centroid and dominant frequency of various respiratory disease conditions, including COVID-19

| **Disease Conditions** | **ZCR Mean values** | **Spectral centroid mean values** | **Dominant Frequency mean values** |
| --- | --- | --- | --- |
| Normal | 0.168 | 2249 | 844 |
| ILD | 0.099 | 2053 | 436 |
| COPD | 0.08 | 1947 | 393 |
| Asthma | 0.112 | 2093 | 528 |
| Pneumonia | 0.118 | 2249 | 546 |
| COVID-19 | 0.246 | 3710 | 1287 |
| COVID-19 (Low severity) | 0.203 | 3300 | 891 |

### LIME data comparison

Extremely low spectral frequencies have been observed in conditions such as Normal and Pneumonia as compared to asthma, which has an intermediate spectral frequency. On the other hand, we found that the spectral components are very high in diseases in which mucus accumulates in the airways and fluid accumulates in the parenchyma region. High spectral content is the distinguishing feature of COVID-19 cough from other respiratory diseases.

Feature analysis studies of cough sounds have revealed that it could be utilized for distinguishing diseases. The cough duration and frequency distribution has been found to be unique in a specific respiratory disease, including COVID-19 [30, 31].

We compared the LIME maps of various respiratory diseases with COVID-19 which are enlisted in Table 6. It can be seen in the maps that each disease has a unique frequency distribution. Green patches were more dominant for COVID-19 in high frequency regions. Whereas LIME maps for normal subjects were reacting negatively even though it has some green patches present. Similarly, pneumonia maps were also present in the high frequency range but has a stronger predominance in the medium frequency range. Even for Asthma most of the dominant green patches were seen in the medium frequency region.

**Table 6:**
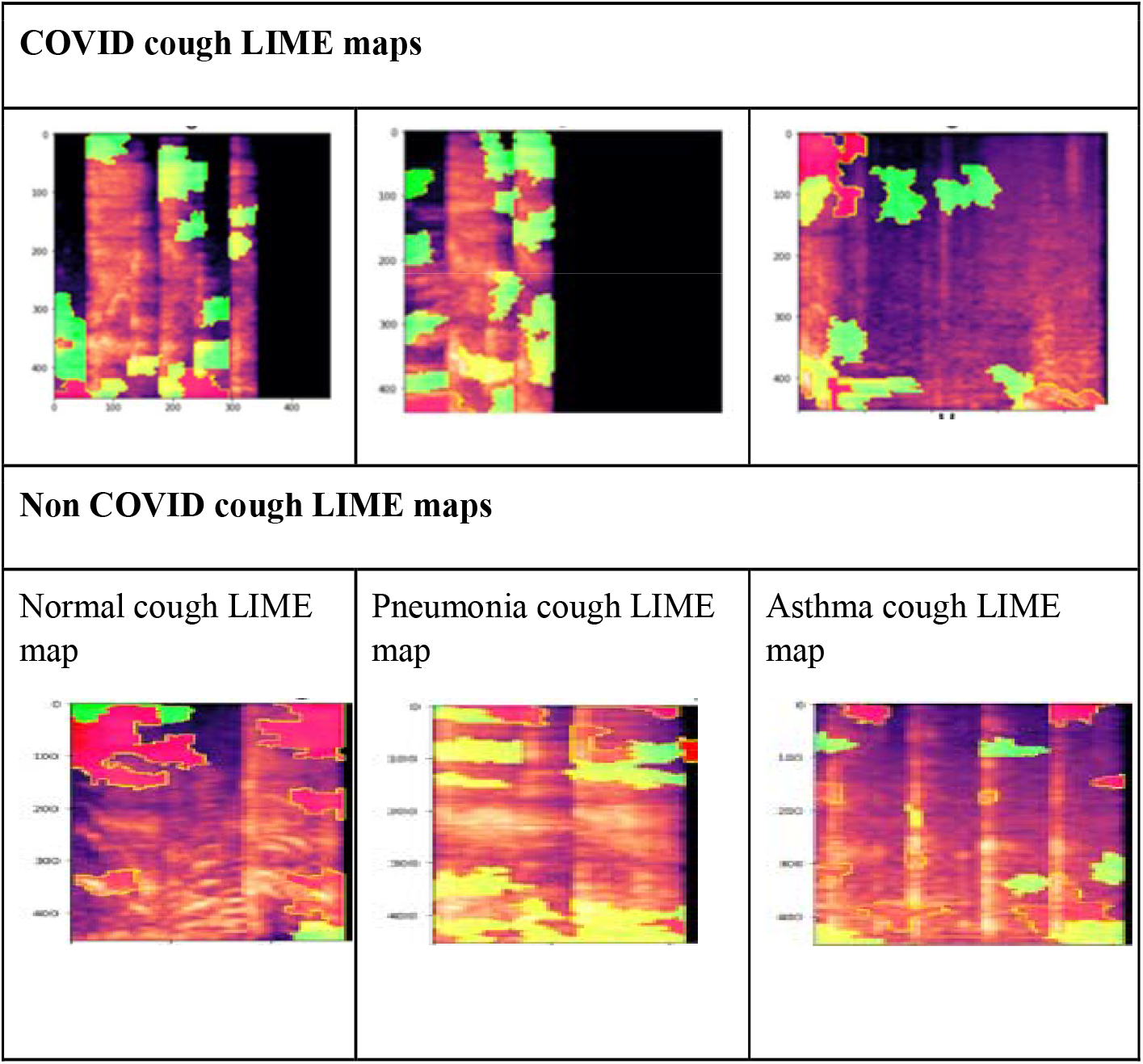
List of different respiratory diseases showing characteristic cough signature, cough spectrograms and related LIME maps

From the LIME map analysis, we can conclude that COVID-19 related cough has a unique signature. These key signatures are detected by features extracted from coughs that can be further identified and characterized by machine learning models.

### Performance Parameters of Model in Validation Phase

Out of 234 subjects participated in the validation phase, 22 were found to be COVID-19 positive and 211 COVID-19 negative by standard diagnostic methods such as RT-PCR.

Results of 1 subject remained inconclusive, hence didn’t consider that datapoint. In this phase only one cough record was collected from each subject. Confusion matrix for validation phase of Swaasa model is illustrated in Table 7, where the row represents the actual label, and the column represents predicted label. An accuracy of 75.54% with 95.45% sensitivity and 73.46% specificity was achieved for the Validation phase (Table 8).

**Table 7:** Confusion matrix for the validation phase

|  | COVID-19 Likely - Yes | COVID-19 Likely - No |
| --- | --- | --- |
| COVID-19 Likely - Yes | 21 (TP) | 1 (FN) |
| COVID-19 Likely - No | 56 (FP) | 155 (TN) |

**Table 8:**
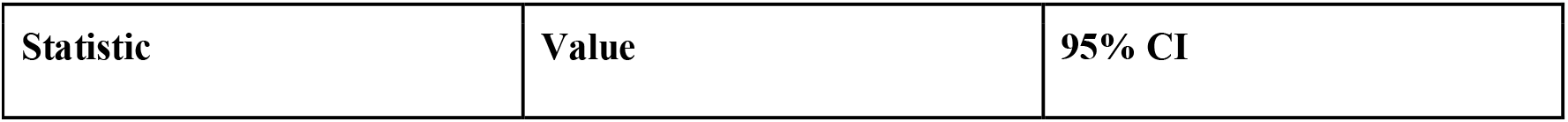
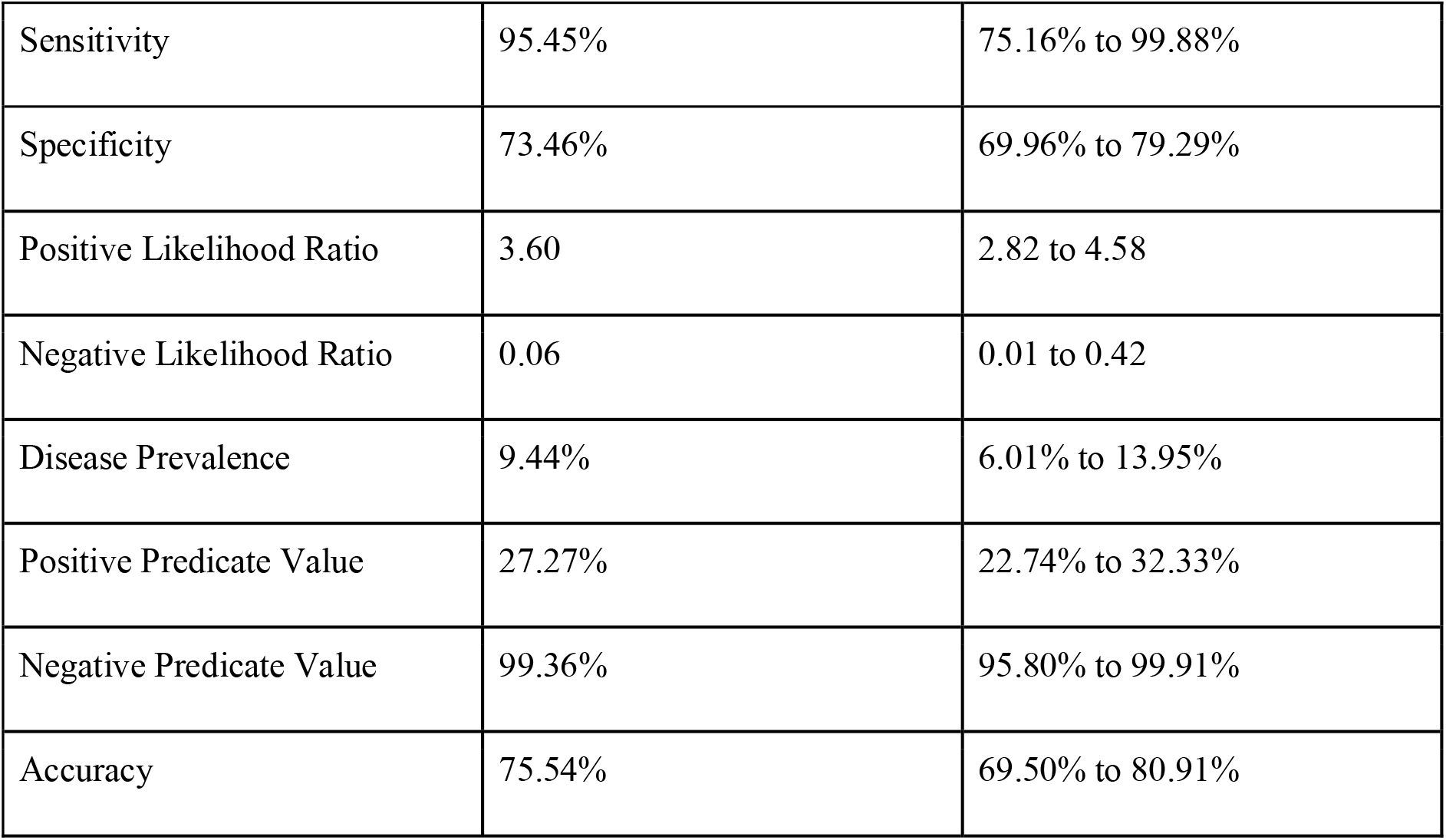
Performance metrics of the validation phase

| Statistic | Value | 95% CI |
| --- | --- | --- |
| Sensitivity | 95.45% | 75.16% to 99.88% |
| Specificity | 73.46% | 69.96% to 79.29% |
| Positive Likelihood Ratio | 3.60 | 2.82 to 4.58 |
| Negative Likelihood Ratio | 0.06 | 0.01 to 0.42 |
| Disease Prevalence | 9.44% | 6.01% to 13.95% |
| Positive Predicate Value | 27.27% | 22.74% to 32.33% |
| Negative Predicate Value | 99.36% | 95.80% to 99.91% |
| Accuracy | 75.54% | 69.50% to 80.91% |

### Model Output in the Pilot phase

A total of 183 patients were recruited for the pilot testing phage. Out of these 183 subjects Swaasa was able to identify 82 subjects as having a likely presence of SARS-CoV-2. Out of these 82 subjects, 58 truly turned out to be COVID-19 positive with a Positive predictive value (PPV) of 70.73%. The confusion matrix for this phase is enlisted in Table 9.

**Table 9:** Confusion matrix for the pilot phase

|  | COVID-19 Likely - Yes | COVID-19 Likely - No |
| --- | --- | --- |
| COVID-19 Likely - Yes | 58 (TP) | 47 (FN) |
| COVID-19 Likely - No | 24 (FP) | 48 (TN) |

The screening of COVID-19 patients by Swaasa is time saving as compared to the currently used traditional procedures. Additionally, Swaasa does not need any trained professional; a community healthcare worker can also perform the screening. The technician did not need any specialised equipment or supplies. The only prerequisites to complete the exam are a smartphone and a stable internet connection.

## Discussion

Accelerated research to find the origin, cause and cure for SARS-CoV-2 infection has resulted in controlling the outspread of COVID-19 to some extent [32]. However, the excessive cost of rapid screening diagnostic kits as well as multiple genetic variants of the virus poses a major hindrance in conducting large scale screening operations [33]. Various researchers across the globe are working to find a cost-effective solution to keep a check on the rapidly mutating virus [34, 35].

Machine learning (ML) has immense potential for accurate and rapid detection of various medical conditions [36], including COVID-19, using computed tomography (CT), chest radiography (CXR), and even coughing pattern [37, 38]. Numerous studies have been conducted in the past to use the information in the cough sound to diagnose and predict the outcome of various diseases, including lung cancer, bronchitis, pneumonia, COPD, and asthma [39, 40].

In a similar study the authors extracted the MFCCs from cough recordings and fed them into a pretrained CNN model, which resulted in an AUC of 97% with a sensitivity and a specificity of 94.2% [17]. Another AI-based COVID-19 cough classifier study includes the analysis of cough recorded over a smartphone, which was able to distinguish COVID-19 positive cases from both COVID-19 negative and healthy coughs. An AUC of 98% was achieved using the Resnet50 classifier to discriminate between COVID-19 positive and healthy coughs, while to differentiate between COVID-19 positive and negative coughs an LSTM classifier was used with an AUC of 94% [19]. In one of the studies both coughs and breathing sounds were used to identify how distinct COVID-19 sounds were as compared to asthma patients or healthy individuals. They highlighted that how a simple binary machine-learning classifier can distinguish COVID-19 cough sounds from healthy subjects by achieving an overall AUC of above 80% [41]. A recent study made use of an ensemble-based multi-criteria decision-making (MCDM) method for detecting COVID-19 from cough sound data and achieved an AUC of 95% [38].

In our study, we developed the Swaasa AI platform by merging the final output layers of the two separate models i.e., the tabular model (training input: primary and secondary features) and CNN model (training input: MFCC spectrograms) because it provides better prediction outcome as compared to the either logical repression or CNN model used alone by other researchers [19, 38, 41]. We conducted the derivation phase, validation phase and pilot screening on a comparatively large cohort, whereas previous studies were performed on either smaller scale or crowdsource datasets which are highly unreliable [38].

Our model achieved an accuracy of 75.54% with 95.45% sensitivity and 73.46% specificity in the clinical validation phase. The pilot testing was undertaken in a real primary care setting to test the accuracy of the tool. Upon deployed as a screening and triaging tool prior to molecular testing, Swaasa was proven statistically effective in prioritizing at-risk patients for confirmatory testing. In the pilot phase also, the model achieved a positive prediction value of 55.24% in a clinical setup at a tertiary care hospital.

Comparing the performance of current rapid diagnostic tests available for COVID-19, Swaasa’s technical and clinical validation results indicate that the device is primarily intended to be used as a screening tool and could be utilized to prioritize the at-risk COVID-19 patients for further evaluation.

The pandemic will truly be over only when the entire world’s population is vaccinated. In this context, the primacy of access to testing at scale, with instantaneous results becomes key. With its frugal operation mode, the Swaasa AI Platform holds out the promise of filling the grossly unmet need of ubiquitous, cost-effective, instantaneous testing in inaccessible, resource-poor parts of the world.

## Data Availability

All data produced in the present study are available upon reasonable request to the authors

## Acknowledgement

This study is supported by the UK Government (British High Commission, New Delhi). This is a commissioned research report on commercial terms between C-CAMP and the UK Government (British High Commission, New Delhi). We would also like to acknowledge the team from Andhra Medical College Visakhapatnam for all the support provided

## Data availability

Due to the nature of this research, participants of this study did not agree for their data to be shared publicly. However, the detailed analysis can be shared upon reasonable request.

## Author contributions

PL and DM defined study protocol, including the study design and methodology. NR conceptualized the idea of using cough sounds for screening and diagnosing COVID-19. GR performed literature review and data analysis. BM, CJ, SDP and NKB were involved in device development. VY created value proposition for the device. SS assisted in executing the project at AMC by providing all the resources and extending research capabilities. CG and GR performed data analysis, sample size estimation and result analysis. KKP provided subject matter expertise. GR and PF wrote the manuscript. All the authors provided intellectual inputs and helped in preparing the manuscript.

